# ICU patients with influenza during the 2025–26 season: a French prospective nationwide cohort study

**DOI:** 10.64898/2026.02.20.26346693

**Authors:** Nicolas de Prost, Pierre Bay, Marie Le Goff, Sébastien Préau, Aurélie Guigon, François M Beloncle, Caroline Lefeuvre, Anaïs Dartevel, Sylvie Larrat, Rémi Coudroy, Luc Deroche, Cédric Darreau, Jean Thomin, Cécile Aubron, Adissa Tran, Fabrice Uhel, Quentin Le Hingrat, Fabienne Tamion, Alice Moisan, Antoine Guillon, Lynda Handala, Bertrand Souweine, Cecile Henquell, Kada Klouche, Edouard Tuaillon, Charles Damoisel, Anne-Marie Roque-Afonso, Elyanne Gault, Pierre Cappy, Alexandre Soulier, Jean-Michel Pawlotsky, Frédéric Lemoine, Marie-Anne Rameix-Welti, Etienne Audureau, Slim Fourati, the SEVARVIR investigators

## Abstract

**Importance:** Recent reports have highlighted an intense influenza activity related to the circulation of the influenza A(H3N2) subclade k variant. There is no data available on the impact of the emergence of H3N2 subclade k on the severity of the 2025–2026 epidemic or on the clinical phenotype of patients requiring admission to the intensive care unit (ICU).

**Objective:** To compare the clinical presentation, hospital mortality and virological characteristics of patients with laboratory-confirmed influenza infection included in French intensive care units during the 2025–2026 epidemic season with those of patients admitted during the 2024–2025 season. We also aimed at measuring the impact of the A(H3N2) subtype on hospital mortality during the 2025-2026 season.

**Design:** Prospective, multicenter, observational SEVARVIR cohort study including patients admitted during the 2024-2025 and 2025-2025 influenza seasons.

**Setting:** Forty-two French ICUs

**Participants:** Adult patients with laboratory-confirmed influenza infection

**Interventions:** none

**Main Outcomes and Measures:** The primary outcome measure was in-hospital mortality.

**Results:** Patients admitted in intensive care units for influenza in 2024-2025 (n=360) and 2025-2026 (n=325) were included in the French nationwide prospective multicentre SEVARVIR study. There was no significant difference in day-28 mortality between the seasons (12.7%, n=45/355 vs 16.5% n=28/170; p=0.28). In the 2025-26 season, 49% had the A(H1N1) subtype and 51% the A(H3N2) subtype (k subclade: 77%). The univariable Cox analysis revealed that patients infected with A(H3N2) viruses were at greater risk of death over time. Multivariable Cox analysis revealed that during the 2025-2026 season, age (adjusted hazard ratio, aHR=1.05 [1.00;1.11]; p=0.046) and the clinical frailty scale (aHR=1.82 [1.26;2.72]; p=0.001) were associated with an increased risk of death. The A(H3N2) subtype was not associated with an increased risk of death (aHR=1.13 [0.32;4.51]; p=0.85). Phylogenetic analyses from our ICU cohort together with 300 contextual sequences from community-acquired influenza cases collected during the same period showed no clustering according to severity.

**Conclusions and Relevance:** This French national prospective observational study, found that the emergence of the influenza A(H3N2) subclade K was associated with an increased risk of death in univariable but not multivariable analysis, adjusting for host-related factors.

**Trial Registration:** NCT051625

**Key Points:** **Question:** What impact did the 2025–26 influenza epidemic and the A(H3N2) variant have on the mortality of patients admitted to intensive care units?

**Findings:** In this prospective, nationwide cohort study of 685 patients admitted to intensive care units with severe influenza during the 2024–25 or 2025–26 seasons, no difference in hospital mortality was observed between the two seasons. Patients infected with the A(H3N2) virus, 77% of which corresponded to clade k, were at higher risk of death in univariable but not in multivariable analysis after adjusting for age and clinical frailty scale.

**Meaning:** Patients in intensive care units with severe A(H3N2) infection during the 2025/2026 season were not at higher risk of death after adjusting for confounding variables.

## Introduction

Recent reports have highlighted an intense influenza activity related to the circulation of the influenza A(H3N2) subclade k variant. The unusually protracted Australian influenza season, which was mainly caused by a mix of influenza A (H1N1) and A(H3N2) viruses, was due to a late circulation of the new influenza A(H3N2) variant of subclade K (J.2.4.1) with multiple hemagglutinin (HA) substitutions ^1^. Reports then emerged of an early and severe influenza circulation in Japan and the UK caused by the A(H3N2) subclade K ^2^. Data from the Centers for Disease Control and Prevention (CDC) confirmed the impact of the epidemic with 280,000 hospitalizations and 12,000 deaths recorded in the USA by 31 January 2026 ^3^. In Europe, weekly reports from the European Centre for Disease Control (ECDC) showed that the influenza epidemic peaked in week 52 ^4^. To our knowledge, there is no data available on the impact of the emergence of H3N2 subclade k on the severity of the 2025–2026 epidemic or on the clinical phenotype of patients requiring admission to the intensive care unit (ICU). The SEVARVIR study is a French prospective multicentre observational cohort study including patients requiring admission to the ICU for acute hypoxemic respiratory failure (AHRF) with a laboratory-confirmed respiratory viral infection. In order to evaluate the clinical significance of this emergence, we compared data from patients with laboratory-confirmed influenza infection who were included in the SEVARVIR study during the 2025–2026 epidemic season with data from patients admitted during the 2024–2025 season.

## Methods

### Patients and clinical data

This is a sub-study of the prospective, multicenter observational SEVARVIR cohort study ^5–8^. Patients admitted to one of the 42 participating ICUs (see **eTable 1** in supplement for the list of participating centers) between December 7th, 2021 and February, 11th 2026 were eligible for inclusion in the SEVARVIR cohort study (NCT05162508) if they met the following inclusion criteria: age ≥18 years, infection by a respiratory virus confirmed by a positive reverse transcriptase-polymerase chain reaction (RT-PCR) in nasopharyngeal swab samples, or any other respiratory samples (e.g., tracheal aspirate, broncho-alveolar lavage fluid), admission to the ICU for acute respiratory failure (i.e., peripheral oxygen saturation (SpO2) ≤90% and need for supplemental oxygen or any kind of ventilator support), patient or next of kin informed of study inclusion. Patients with respiratory virus infection but no acute respiratory failure were not included. For this sub-study focused on severe influenza, we included patients who had laboratory-confirmed influenza infection (i.e., positive influenza PCR).

Demographic, clinical and laboratory variables were recorded upon ICU admission and during their stay in the ICU. Patients’ frailty was assessed using the Clinical Frailty Scale ^9^. The disease severity at ICU admission was evaluated using the World Health Organization (WHO) 10-point ordinal scale^10^, the sequential organ failure assessment (SOFA) score ^11^, and the simplified acute physiology score (SAPS) II score ^12^.

### Ethical approval

The study was approved by the *Comité de Protection des Personnes Sud-Méditerranée* I (N° EudraCT/ID-RCB: 2021-A02914-37). Informed consent was obtained from all patients or their relatives. The study was conducted in accordance with the 1964 Helsinki Declaration and subsequent amendments.

### Influenza sequencing

Full-length influenza A and B virus genomes (8 segments) were obtained using a whole-genome amplification strategy based on the French National Influenza Reference Center protocol (see additional Methods in the supplement).

### Phylogenetic analyses

We randomly selected 300 high-quality French contextual samples of each subtype from the National Reference Center (spanning the 2024–2025 and 2025–2026 seasons) using goalign subset (version devd0f3e37) ^13^. SEVARVIR sequences containing >20% ambiguous nucleotides (N) were filtered out using goalign clean ^13^, and those derived from samples with high CT values (≥30) were excluded. All sequences were annotated with Nextclade v3.18.0 ^14^ and aligned using Augur v32.1.0 ^15^ (see additional Methods in the supplement).

### End Points

The primary end point was in-hospital mortality. The secondary end points included day-28 mortality, as well as the duration of ICU stay.

### Statistical analyses

Descriptive results are presented as means (± standard deviation [SD]) or medians (1st–3rd quartiles) for continuous variables, and as numbers with percentages for categorical variables. Two-tailed p values_<_0.05 were considered statistically significant. Unadjusted comparisons according to influenza season status (2024-25 season vs. 2025-26 season) and influenza subtype (A(H1N1)pdm09 vs. A(H3N2)) for the 2025-26 season were performed using Chi-square or Fisher’s exact tests for categorical variables, and t-tests or Mann–Whitney tests for continuous variables, as appropriate. Exploratory unadjusted comparisons according to influenza subtype (A(H1N1)pdm09 vs. A(H3N2) vs. subtype undetermined vs. respiratory sample unavailable for sequencing) for the 2025-26 season 2025-26 were performed using Chi square or Fisher’s exact tests for categorical variables, and ANOVA or Kruskal–Wallis tests for continuous variables, as appropriate. Missing data were not imputated. Analyses of the association between the influenza subtype during the 2025-26 season (i.e., A(H1N1)pdm09, A(H3N2), undetermined subtype) and mortality relied on univariable and multivariable Cox models. Variables associated with a *p*_<_0.20 in univariable analysis were entered in the multivariable model, and a stepwise backward approach was then applyied by retaining only the variables which were statistically significant at the *p*_<_0.05 level, as well as those previously determined to be important confounding factors. Adjusted hazard ratios (aHRs) along with their 95% confidence intervals (CI) were computed. Unadjusted and adjusted survival analyses were performed to assess the prognostic value of the infecting subtype during the season 2025-26 on day-28 survival. Day-28 survival curves were plotted by the Kaplan-Meier method, using log-rank tests to assess significance for between-group comparison.

To illustrate differences in phenotypes of patients according to infecting influenza virus subtype status, we performed an exploratory unsupervised clustering analysis using the Kohonen’s self-organized map (SOM) methodology ^16^, allowing us to build 2-dimensional maps from multidimensional datasets (see additional Methods in the supplement).

All analyses involving survival and the infecting influenza virus subtype during the 2025–26 season (i.e., Cox models, SOM clustering analysis) were performed only among patients with available follow-up of at least 30 days and for whom a respiratory sample had been sequenced (*N*=158).

## Results

### Seasonal comparison: 2025–2026 vs. 2024–2025

A total of 325 patients admitted to the ICU with AHRF associated with an influenza infection were included in the SEVARVIR study during the 2025–26 season, compared to 360 patients during the 2024-25 season (**Figures 1A and 1B**). Subtype identification was available for 144 (44.3%) of the 325 patients of the 2025-26 season, including 70 (48.6%) infected with A(H1N1)pdm09 and 74 (51.4%) infected with A(H3N2). A significantly higher proportion of patients were infected with A(H3N2) in the 2025-26 season than in the 2024–25 season (*N*=86/360, 39.6%, p<0.001, **Table 1**). Patients admitted during the 2025–26 season were significantly older than those admitted during the previous season with a median age of 67.9 years [59.0;74.8] vs. 65.2 years [54.8;72.2] (p=0.009). There was a high prevalence of cardiovascular comorbidities, with 20% of patients being immunocompromised, without significant differences between the two seasons. The influenza vaccination coverage rate was 23.5% during the 2024-25 season and 28.4% during the 2025-26 season (p=0.24). Although the proportion of patients requiring invasive mechanical ventilation (IMV) upon admission to the ICU did not differ significantly between seasons, patients admitted during the 2025–26 season had a less severe presentation. They had significantly lower arterial lactate levels (1.5 mmol/L [1.1;2.4] vs. 1.7 mmol/L [1.2;2.9], p=0.049) and tended to have lower SOFA scores. During their stay in the ICU, patients admitted during the 2025–26 season required vasopressors significantly less frequently (p=0.049) and were less likely to require IMV than those admitted during the previous season. During the 2025–26 season, almost all patients received oseltamivir, significantly more frequently than during the previous season (93.9% vs 86.0%; p=0.002). By contrast, corticosteroids were prescribed significantly less frequently to patients admitted during the 2025–26 season (33.3% vs 43.4%; p=0.015).

**Figure 1.**
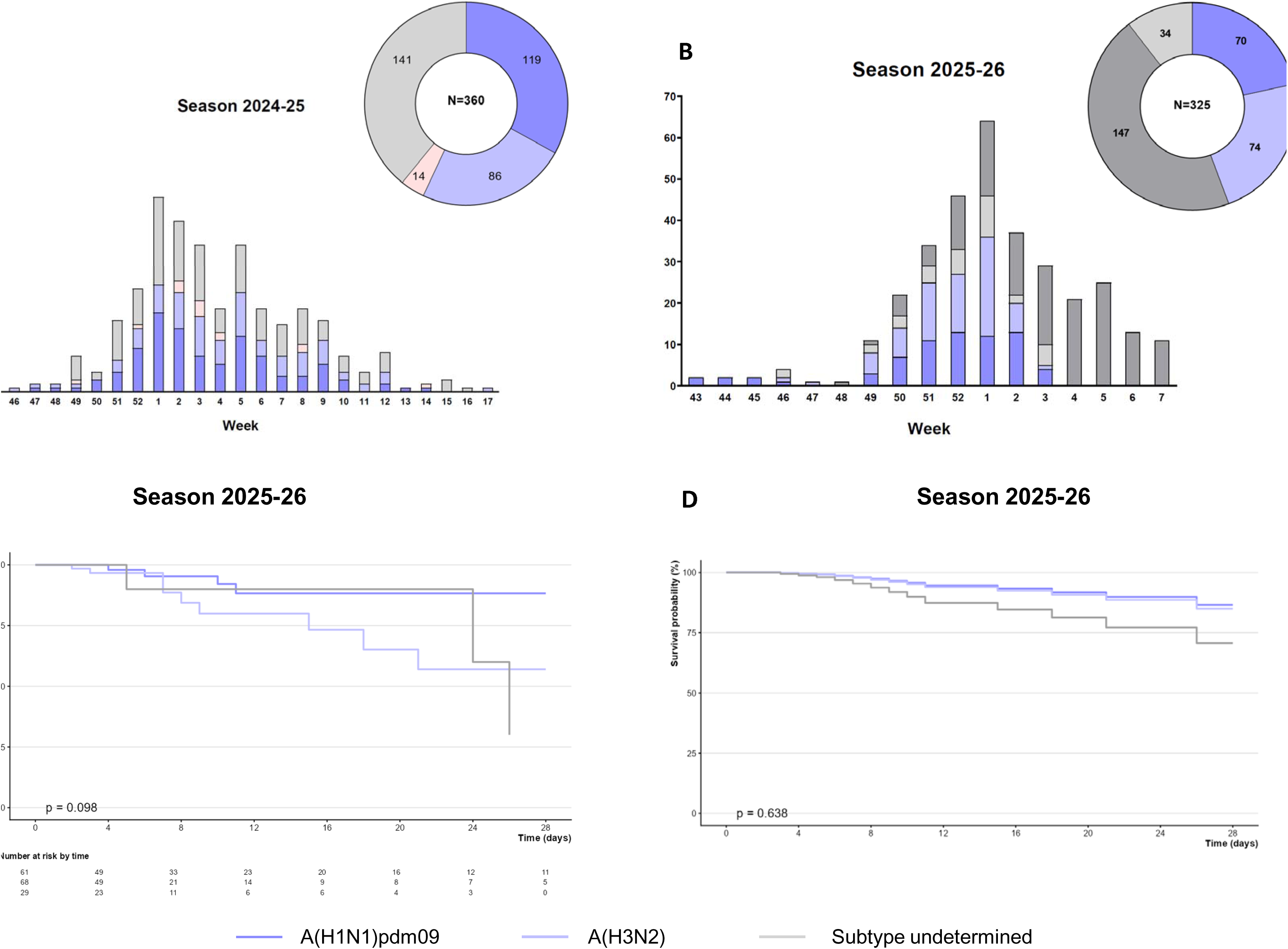
Seasonal distribution of ICU admissions and day-28 mortality according to influenza virus subtype, France, 2024–26. Number of critically ill patients with influenza included in the study each week during the 2024–25 season (Panel A) and 2025-26 season (Panel B), stratified by influenza virus subtype. The histogram shows the number of inclusions by calendar week. The pie chart summarises the overall distribution of influenza virus subtypes throughout the season. Kaplan–Meier survival analysis curves for 28-day mortality according to the influenza subtype. Panel C shows the unadjusted survival Kaplan–Meier and Panel D shows the Cox-adjusted survival curves. Numbers of patients at risk and number of events are shown in the risk table below the graph. P value come from the log-rank test. Patients infected with A(H1N1) are shown in dark blue, patients infected with A(H3N2) in light blue, those infected with influenza B in pink, and patients with unknown subtypes or samples unavailable for sequencing are shown in light grey and dark grey, respectively. Note that patients with samples that were unavailable for sequencing were not considered in the survival analyses in panels C and D.

**Table 1.** Characteristics of the 685 critically ill patients infected with influenza according to the season.

| Variable | N | 2024-2025,<br>n=360 | 2025-2026,<br>n=325 | p |
| --- | --- | --- | --- | --- |
| **Demographics and comorbidities** |  |  |  |  |
| Sex, females | 685 | 153 (42.5) | 130 (40.0) | 0.53 |
| Age, years | 685 | 65.2 [54.8;72.2] | 67.9 [59.0;74.8] | **0.009** |
| Diabetes | 520 | 86 (29.7) | 75 (32.6) | 0.50 |
| Obesity | 671 | 101 (28.6) | 91 (28.6) | 0.36 |
| Chronic heart failure | 519 | 60 (20.7) | 40 (17.5) | 0.37 |
| Hypertension | 520 | 156 (53.8) | 125 (54.3) | 0.93 |
| Chronic respiratory failure | 519 | 107 (37.0) | 90 (39.1) | 0.65 |
| Chronic renal failure | 520 | 46 (15.9) | 31 (13.5) | 0.46 |
| Cirrhosis | 519 | 4 (1.4) | 1 (0.4) | 0.39 |
| Immunosuppression | 520 | 64 (22.1) | 46 (20.0) | 0.59 |
| Immunosuppression category | 503 |  |  | 0.70 |
| None |  | 226 (79.9) | 184 (83.6) |  |
| Solid organ transplant |  | 14 (4.9) | 7 (3.2) |  |
| Onco-hematological malignancies |  | 21 (7.4) | 14 (6.4) |  |
| Others<sup>1</sup> |  | 22 (7.8) | 15 (6.8) |  |
| Number of comorbidities | 520 | 2.0 [1.0;3.0] | 2.0 [1.0;3.0] | 0.76 |
| Clinical frailty scale | 609 | 3.0 [2.0;4.0] | 3.0 [2.0;4.0] | 0.92 |
| **Influenza infection and Vaccination** |  |  |  |  |
| Previous Influenza infection | 465 | 24 (9.4) | 32 (15.3) | 0.062 |
| Influenza vaccination | 465 | 58 (23.5) | 62 (28.4) | 0.24 |
| Influenza RNA detection in nasopharyngeal swabs, Ct | 502 | 35.0 [27.0;40.0] | 31.0 [26.0;39.0] | **0.023** |
| *Influenza subtype* | 502 |  |  | **<0.001** |
| H1N1 |  | 119 (36.7) | 70 (39.3) |  |
| H3N2 |  | 86 (26.5) | 74 (41.6) |  |
| Influenza B |  | 14 (4.3) | 0 (0.0) |  |
| Undetermined subtype |  | 105 (32.4) | 34 (19.1) |  |
| Unavailable for sequencing | 685 | 36 (10.0) | 147 (45.2) | **<0.001** |
| **Patients severity upon ICU admission and biological features** |  |  |  |  |
| SAPS II score | 572 | 39.5 [30.0;50.5] | 38.0 [29.0;49.5] | 0.40 |
| SOFA score | 618 | 5.0 [3.0;9.0] | 4.0 [3.0;8.0] | 0.057 |
| PaO<sub>2</sub>/FiO<sub>2</sub> ratio, mmHg | 615 | 156 [102;227] | 163 [105;240] | 0.14 |
| Arterial lactate levels, mM | 597 | 1.70 [1.20;2.90] | 1.50 [1.10;2.40] | **0.049** |
| Blood leukocytes, G/L | 637 | 8.6 [5.7;14.0] | 8.9 [5.6;13.0] | 0.74 |
| Blood lymphocytes, G/L | 531 | 0.70 [0.40;1.20] | 0.70 [0.40;1.10] | 0.95 |
| Blood platelets, G/L | 636 | 187 [130;248] | 190 [133;246] | 0.45 |
| Serum urea levels, mM | 637 | 8.0 [5.0;14.0] | 7.2 [5.0;12.6] | 0.14 |
| Serum creatinine levels, µM | 644 | 86.0 [62.0;150] | 84.0 [60.0;135] | 0.57 |
| *Oxygen/ventilatory support* | 643 |  |  | 0.67 |
| Oxygen |  | 67 (18.7) | 64 (22.5) |  |
| High flow oxygen |  | 98 (27.4) | 74 (26.0) |  |
| NIV/C-PAP |  | 68 (19.0) | 55 (19.3) |  |
| Invasive MV |  | 125 (34.9) | 92 (32.3) |  |
| Vasopressor support | 615 | 115 (33.6) | 80 (29.3) | 0.26 |
| Bacterial coinfection | 641 | 124 (34.5) | 82 (29.1) | 0.15 |
| **Management during ICU stay** |  |  |  |  |
| Invasive MV | 646 | 158 (44.0) | 106 (36.9) | 0.076 |
| Prone positioning | 599 | 61 (18.0) | 41 (15.7) | 0.51 |
| MV duration, days* | 221 | 9.0 [5.0;22.0] | 9.5 [5.0;17.0] | 0.65 |
| Live-ventilator-free days at day-28* | 524 | 28.0 [11.0;28.0] | 28.0 [13.0;28.0] | 0.70 |
| Vasopressor support | 643 | 146 (40.7) | 94 (33.1) | **0.049** |
| Duration of vasopressors, days* | 211 | 4.0 [2.0;12.0] | 43.50 [1.0;10.0] | 0.53 |
| Renal replacement therapy | 638 | 41 (11.5) | 31 (11.1) | 0.88 |
| Ventilator-acquired pneumonia*<sup>2</sup> | 229 | 46 (29.5) | 19 (26.0) | 0.64 |
| Oseltamivir | 566 | 276 (86.0) | 230 (93.9) | **0.002** |
| Corticosteroids | 563 | 139 (43.4) | 81 (33.3) | **0.018** |
| **Outcomes** |  |  |  |  |
| Day-28 mortality* | 524 | 45 (12.7) | 28 (16.5) | 0.28 |
| Duration of ICU stay, days* | 558 | 7.0 [4.0;15.0] | 6.0 [3.0;11.0] | 0.051 |

Results are N (%), means (±standard deviation) or medians (interquartile range, i.e., quartile 1; quartile 3).
ICU intensive care unit, Ct cycle threshold, SOFA Sequential Organ Failure Assessment, SAPS II Simplified Acute Physiology Score II, NIV non-invasive ventilation, C-PAP continuous-positive airway pressure, MV mechanical ventilation.
<sup>1</sup>Includes HIV infection, long-term corticosteroid treatment, and other immunosuppressive treatments.
<sup>2</sup>VAP episodes were recorded per definition in patients under IMV since more than 48h.
\*These data are reported only for patients with minimum potential follow-up of 30 days between inclusion and the data cut-off date (i.e. Feb 11<sup>th</sup> 2026; 2024–25 season: N=357; 2025–26 season: N=209).
Two-tailed p-values come from unadjusted comparisons using Chi-square or Fisher's exact tests for categorical variables, and t-tests or Mann–Whitney tests for continuous variables, as appropriate. No adjustment for multiple comparisons was performed. Bolded p-values are significant at the p<0.05 level.

### 2025–2026 season: A(H1N1)pdm09 vs. A(H3N2)

During the 2025–26 season, respiratory samples from 147 out of 325 patients were unavailable for influenza virus sequencing. Of the patients for whom a respiratory sample was available, 70 (39.3%) were infected with influenza A(H1N1)pdm09, 74 (41.6%) with influenza A(H3N2), and the subtype was undetermined after PCR-specific subtyping for 34 (19.1%) patients because of low viral load (Ct>38) (**eTable 2** in the supplement).

Patients infected with A(H3N2) were significantly older than those infected with A(H1N1) pdm09 (median ages 67.9 [59.0-74.9] vs 65.2 years [54.8-72.2], p=0.014) (**Table 2**). Influenza vaccination coverage was 20% in the A(H1N1) pdm09 group and 29% in the A(H3N2) group (p=0.43). There were no significant differences in comorbidities between the two groups, except for a significantly higher prevalence of hypertension among patients infected with the A(H3N2) subtype. Upon admission to the ICU, patients infected with the A(H1N1)pdm09 subtype had lower PaO_O_/FiO_O_ ratios than patients infected with the A(H3N2) subtype (156 mmHg [102;227] vs 169 mmHg [108;242], p=0.009). During their stay in the ICU, patients infected with A(H1N1)pdm09 tended to require more organ support than those infected with A(H3N2), including IMV, vasopressors, prone positioning, and renal replacement therapy.

**Table 2.** Characteristics of the 144 critically ill patients infected with influenza and available subtype according to the infecting subtype during 2025-2026 season.

| Variable | N | H1N1<br>N=70 | H3N2<br>N=74 | p |
| --- | --- | --- | --- | --- |
| **Demographics and comorbidities** |  |  |  |  |
| Sex, females | 144 | 27 (38.6) | 29 (39.2) | >0.99 |
| Age, years | 144 | 66.6 [56.5;74.7] | 72.0 [61.6;77.6] | 0.063 |
| Diabetes | 116 | 18 (34.0) | 24 (38.1) | 0.70 |
| Obesity | 643 | 26 (38.2) | 19 (26.4) | 0.18 |
| Chronic heart failure | 115 | 8 (15.4) | 12 (19.0) | 0.63 |
| Hypertension | 116 | 24 (45.3) | 44 (69.8) | **0.009** |
| Chronic respiratory failure | 116 | 17 (32.1) | 23 (36.5) | 0.70 |
| Chronic renal failure | 116 | 7 (13.2) | 13 (20.6) | 0.33 |
| Cirrhosis | 116 | 0 (0.0) | 1 (1.6) | >0.99 |
| Immunosuppression | 116 | 15 (28.3) | 9 (14.3) | 0.071 |
| Immunosuppression category | 110 |  |  | 0.17 |
| None |  | 38 (77.6) | 54 (88.5) |  |
| Solid organ transplant |  | 1 (2.0) | 3 (4.9) |  |
| Onco-hematological malignancies |  | 5 (10.2) | 2 (3.3) |  |
| Others<sup>1</sup> |  | 5 (10.2) | 2 (3.3) |  |
| Number of comorbidities | 116 | 2.0 [1.0;2.0] | 2.0 [1.0;3.0] | 0.24 |
| Clinical frailty scale | 129 | 3.0 [1.0;4.0] | 3.0 [2.0;4.0] | 0.32 |
| **Influenza infection and Vaccination** |  |  |  |  |
| Previous Influenza infection | 103 | 3 (6.4) | 13 (23.2) | **0.027** |
| Influenza vaccination | 114 | 18 (34.0) | 22 (36.1) | 0.81 |
| Influenza RNA detection in nasopharyngeal swabs, Ct | 144 | 30.0 [25.0;34.0] | 30.0 [25.0;35.0] | 0.87 |
| **Patients severity upon ICU admission and biological features** |  |  |  |  |
| SAPS II score | 131 | 39.5 [29.0;49.0] | 35.0 [25.0;49.0] | 0.43 |
| SOFA score | 137 | 5.0 [3.0;8.5] | 4.0 [3.0;8.0] | 0.18 |
| PaO<sub>2</sub>/FiO<sub>2</sub> ratio, mmHg | 134 | 139 [97.8;223] | 197 [115;280] | 0.11 |
| Arterial lactate levels, mM | 131 | 1.50 [1.10;2.45] | 1.40 [1.00;2.40] | 0.22 |
| Blood leukocytes, G/L | 141 | 8.7 [4.5;13.3] | 8.8 [5.2;10.9] | 0.82 |
| Blood lymphocytes, G/L | 119 | 0.60 [0.40;1.00] | 0.60 [0.40;1.05] | 0.69 |
| Blood platelets, G/L | 141 | 158 [118;242] | 190 [145;231] | 0.16 |
| Serum urea levels, mM | 142 | 7.0 [5.0;13.8] | 8.3 [5.2;12.6] | 0.89 |
| Serum creatinine levels, µM | 142 | 94.0 [58.5;146] | 89.0 [65.0;141] | 0.85 |
| Oxygen/ventilatory support | 142 |  |  | 0.86 |
| Oxygen |  | 15 (22.1) | 19 (25.7) |  |
| High flow oxygen |  | 15 (22.1) | 19 (25.7) |  |
| NIV/C-PAP |  | 14 (20.6) | 14 (18.9) |  |
| Invasive MV |  | 24 (35.3) | 22 (29.7) |  |
| Vasopressor support | 136 | 21 (32.8) | 17 (23.6) | 0.26 |
| Bacterial coinfection | 141 | 15 (22.1) | 17 (23.3) | >0.99 |
| **Management during ICU stay** |  |  |  |  |
| Invasive MV | 142 | 28 (41.2) | 22 (29.7) | 0.16 |
| Prone positioning | 134 | 16 (26.2) | 6 (8.2) | **0.009** |
| MV duration, days* | 38 | 12.0 [9.0;20.0] | 7.0 [3.0;17.0] | 0.22 |
| Live-ventilator-free days at day-28* | 102 | 28.0 [16.0;28.0] | 28.0 [11.0;28.0] | 0.68 |
| Vasopressor support | 141 | 24 (35.8) | 21 (28.4) | 0.37 |
| Duration of vasopressors, days* | 41 | 4.0 [3.0;10.0] | 3.0 [1.0;10.0] | 0.34 |
| Renal replacement therapy | 141 | 11 (16.4) | 5 (6.8) | 0.11 |
| Ventilator-acquired pneumonia*<sup>2</sup> | 129 | 8 (13.1) | 3 (4.4) | 0.11 |
| Oseltamivir | 122 | 53 (93.0) | 62 (95.4) | 0.70 |
| Corticosteroids | 121 | 15 (26.3) | 21 (32.8) | 0.55 |
| **Outcomes** |  |  |  |  |
| Day-28 mortality* | 106 | 4 (7.8) | 11 (20.0) | 0.10 |
| Duration of ICU stay, days* | 124 | 7.0 [4.0;16.0] | 5.0 [3.0;9.0] | 0.065 |

Results are N (%), means ( ± standard deviation) or medians (interquartile range, i.e., quartile 1; quartile 3).
ICU intensive care unit, Ct cycle threshold, WHO World Health Organization, SOFA Sequential Organ Failure Assessment, SAPS II Simplified Acute Physiology Score II, NIV non-invasive ventilation, C-PAP continuous-positive airway pressure, MV mechanical ventilation.
<sup>1</sup>Includes HIV infection, long-term corticosteroid treatment, and other immunosuppressive treatments.
<sup>2</sup>VAP episodes were recorded per definition in patients under IMV since more than 48h.
\*These data are reported only for patients with minimum potential follow-up of 30 days between inclusion and the data cut-off date (i.e. Feb 11<sup>th</sup> 2026; H1N1: N=62; H3N2 season: N=68).
Two-tailed p-values come from unadjusted comparisons using Chi-square or Fisher's exact tests for categorical variables, and t-tests or Mann-Whitney tests for continuous variables, as appropriate. No adjustment for multiple comparisons was performed. Bolded p-values are significant at the $p < 0.05$ level.

To identify subgroups of patients with homogeneous phenotypes, we performed a clustering analysis using the SOM approach. As illustrated in **eFigure 1** in the supplement, this approach revealed polarised distributions of patient characteristics across the maps. Patients who were older and had a higher burden of cardiovascular comorbidities tended to cluster together, as did patients infected with the influenza A(H3N2) subtype. By contrast, patients infected with the influenza A(H1N1)pdm09 subtype were found to cluster with younger patients who had fewer comorbidities, except for immunosuppression, and were characterised by a higher severity of illness on admission to the ICU, as indicated by SOFA and SAPS II scores, PaO_O_/FiO_O_ ratio and arterial lactate levels.

### Factors associated with in-hospital mortality

Univariable and multivariable Cox analyses were performed, among patients with a follow-up period of at least 30 days and a respiratory sample that had been subtyped by centralised subtype specific PCR and sequenced (*N*=158) in order to assess the relationship between viral subtypes and ICU mortality, censored at day 28. The univariable analysis revealed that patients infected with A(H3N2) viruses, as well as older patients, patients with hypertension and patients with an elevated clinical frailty scale or requiring IMV upon ICU admission, were at greater risk of death over time (**eTable 3** in the supplement). Multivariable Cox analysis revealed that age (adjusted hazard ratio, aHR=1.05 [1.00;1.11]; p=0.046) and the clinical frailty scale (aHR=1.82 [1.26;2.72]; p=0.001) were the only variables that remained associated with an increased risk of death, with no statistically significant differences across viral subtypes (global p=0.638) (**eTable 3**). The A(H3N2) subtype was not associated with an increased risk of death (aHR=1.13 [0.32;4.51]; p=0.85). A sensitivity analysis using multivariable Fine-Gray regression modelling accounting for the competing risk of ICU discharge yielded broadly similar results with no differences across viral subtypes (global p=0.94) and associations between increased mortality and increasing age (p=0.077) and clinical frailty scale (p=0.011). Unadjusted Kaplan–Meier curves (**Figure 1C**) and their estimated survival probabilities showed that patients infected with the A(H3N2) subtype tended to have a lower survival probability (p=0.11), but this was not reflected in the Cox model-adjusted survival curves (**Figure 1D**).

### Virological characteristics of influenza A viruses during the 2025–2026 season

Among the 325 patients of the 2025-26 season, the median cycle threshold (Ct) value was 31 [26–39], with no significant difference observed between patients infected with the A(H1N1)pdm09 (n=70) and the A(H3N2) (n=74) subtypes (**Table 2**). Whole-genome sequencing was available for 52 respiratory samples collected during the 2025–26 season, representing 21 A(H1N1)pdm09 and 31 A(H3N2) viruses. All A(H1N1)pdm09 influenza viruses belonged to subclade D.3.1.1, and 77% (*N*=24/31) of A(H3N2) influenza viruses belonged to subclade K.

Phylogenetic analyses of the hemagglutinin gene, including sequences from both the 2024-25 and 2025-26 seasons from our cohort together with 300 contextual sequences from community-acquired, non-severe, influenza cases in France collected during the same period, revealed wide genetic dispersion of sequences, without clustering according to geographical origin, severity of infection (ICU strains vs. community-acquired strains), or 28-day clinical outcome. As expected, sequences segregated predominantly by season (**Figures 2A** and **2B**). This absence of distinct phylogenetic clustering was consistently observed for both A(H1N1)pdm09 and A(H3N2) viruses.

**Figure 2.**
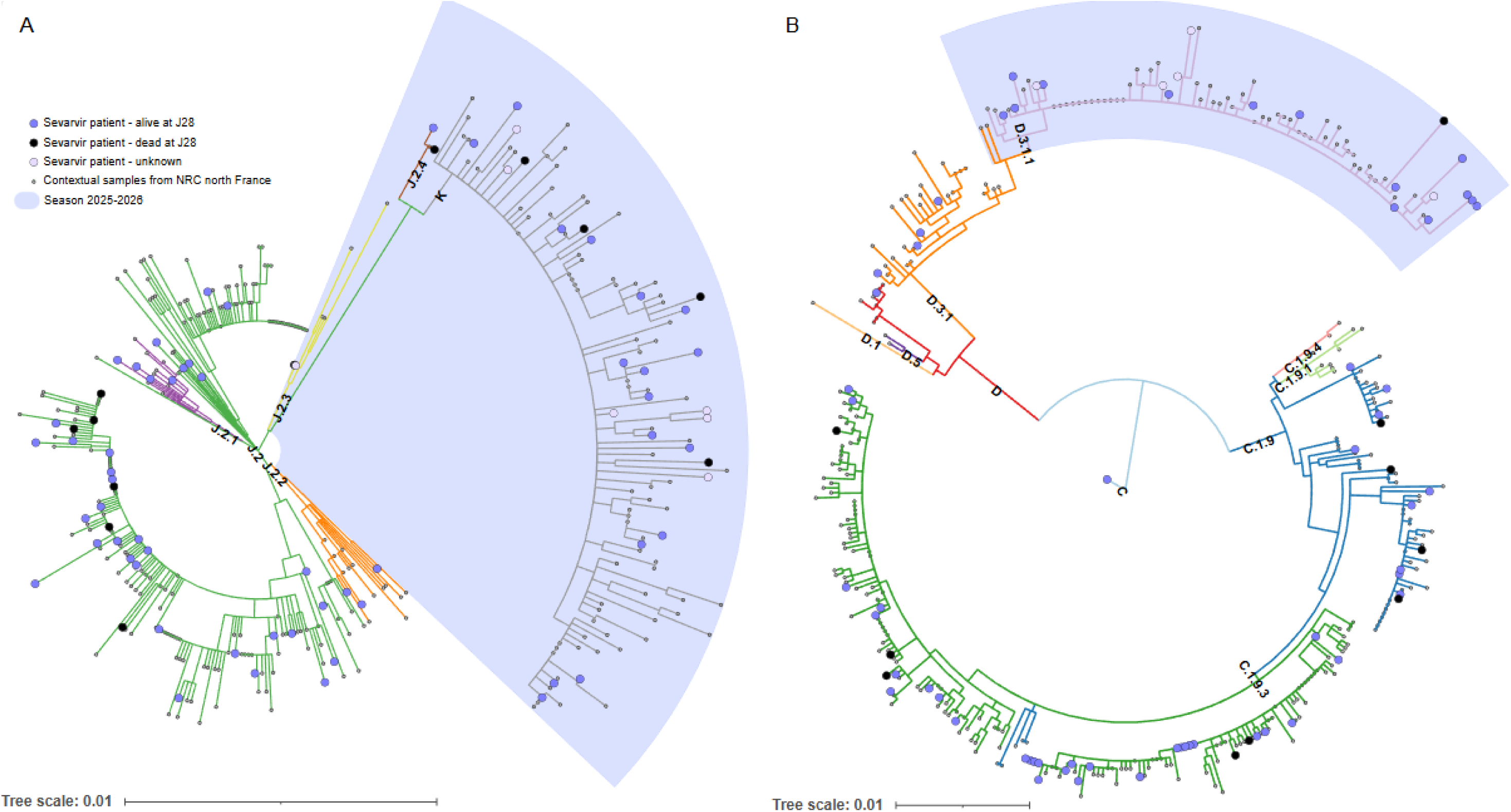
Phylogenetic analysis of SEVARVIR patient-derived influenza sequences in the context of French seasonal epidemics (2024–2025 and 2025–2026, and non-hospitalized cases). (A) H3N2 subtype; (B) H1N1pdm09 subtype; Clades are colour-coded by branch and labelled at their inferred ancestral node. SEVARVIR samples are shown as enlarged circles at terminal nodes, with fill color indicating the patient’s 28-day outcome: black (deceased), blue (survived), or light gray (unknown). Reference sequences from the National Reference Center (NRC) are depicted as smaller circles. Interactive trees are available online for better visualization: A(H1N1): https://itol.embl.de/tree/1579964253244561770829624#, A(H3N2): https://itol.embl.de/tree/15799642539591770831476.

Among patients infected with the A(H1N1)pdm09 subtype, the neuraminidase amino-acid substitution S247N was found in 57% (N=12/21) of the A(H1N1)pdm09 viruses that were sequenced. Exposure to oseltamivir was similar between patients infected with viruses harbouring the NA-S247N substitution (75%, N=9/12) and those infected with viruses lacking this substitution (78%, N=7/9). The NA-S247N substitution was specifically investigated due to its reported association with reduced susceptibility to neuraminidase inhibitors and its recent expansion in Europe, although its relevance remains speculative ^17^.

All 21 patients infected with A(H1N1)pdm09 were either still alive on day 28, or had been discharged from the ICU prior to this date. No difference in 28-day mortality was observed according to NA-S247N status. However, patients infected with NA-S247N viruses were more likely to require IMV than those infected with non-mutated viruses, although this was not statistically significant (33%, n=4/12 vs 11%, n=1/9; p=0.34 by Fisher’s exact test). The combined neuraminidase substitutions NA-I223V and NA-S247N, which were previously shown to reduce oseltamivir susceptibility in vitro ^18^, were observed in a patient. No NA-H275Y substitution was detected among A(H1N1)pdm09 viruses.

## Discussion

To our knowledge, this is the first multicentre study to describe critically ill patients with severe influenza who were admitted to the ICU during the 2025–26 season. This period was characterised by unprecedented global influenza circulation, which was associated with the emergence of the A(H3N2) subclade K variant. This season was marked by intense community transmission and high rates of attendance at the emergency department and hospitalisation ^19^, which were likely driven by the substantial antigenic divergence of the circulating subclade K variant compared with previous subclades ^2,20^. Although epidemiological reports and vaccine effectiveness studies from Australia, the UK, Canada, France, and the United States highlighted the magnitude of this epidemic and suggested moderate overall vaccine effectiveness ^2,20–23^, detailed data on its clinical impact on ICU populations was lacking.

Using data from the SEVARVIR prospective multicentre cohort study, we present the first detailed characterisation of critically ill patients during this atypical season, comparing them with patients admitted during the 2024–25 epidemic. Despite the increase in A(H3N2) circulation in the 2025–26 season, patients admitted during this period appeared to be less severely ill on ICU admission, with lower arterial lactate levels, and tended to require less organ support during their stay in the ICU. These findings are consistent with the global epidemiological data reported by the WHO, which does not indicate an increase in disease severity ^21,24^. Instead, the patients in the 2025–26 cohort were significantly older than those in the 2024-25 cohort. This is consistent with the higher prevalence of the A(H3N2) subtype this year, which is classically associated with elderly populations ^25^. This shift in age distribution is probably due to older individuals being more susceptible in the context of antigenic drift and reduced vaccine effectiveness. Despite these demographic changes, day-28 mortality did not appear substantially different from the previous season. However, focusing on the 2025-26 season, patients infected with the A(H3N2) subtype were found to be at higher risk of death. Yet, after adjusting for important confounders, including age, frailty, and comorbidities, no significant relationship was found between virus subtype and risk of mortality. This suggests that host-related factors (e.g., age and comorbidities in A(H3N2) infections) and intrinsic viral severity ^26,27^ both contribute to clinical outcomes, making it challenging to determine their respective roles in clinical outcomes.

The relative stability of ICU phenotypes and outcomes between seasons and virus subtypes indicate that the public health impact of the subclade K may be primarily due to reduced immunity at the population-level rather than increased intrinsic virulence. Our phylogenetic analyses of the HA gene reinforce this hypothesis, as they showed no specific clustering of ICU-derived sequences compared with community strains, arguing against the existence of a particular viral genetic profile associated with severe disease.

In contrast, we identified frequent neuraminidase polymorphisms among A(H1N1)pdm09 viruses, notably S247N (57%), which was sometimes found alongside I223V (5%) as recently reported in Spain and other regions worldwide ^17^. These substitutions have been reported to reduce susceptibility to oseltamivir ^17,28^, although their clinical relevance remains unclear. Although almost all patients in our cohort received oseltamivir, we could not establish a causal link between these mutations and adverse outcomes. Nevertheless, their presence raises concerns about a potential reduction in the effectiveness of antivirals in cases of severe influenza. This highlights the importance of performing genomic analyses and phenotypic assays alongside clinical data, particularly in ICU settings.

### Strengths and Limitations

Our study has several limitations. First, it was conducted exclusively in France, which may restrict the generalizability of the findings to other countries, particularly those that experienced a more severe influenza season in 2025-26. Second, information on viral subtypes was available for only one third of patients during the 2025–26 season. Third, follow-up is ongoing and therefore outcome data are not yet fully consolidated. Fourth, the assessment of the clinical impact of mutations associated with reduced oseltamivir susceptibility was based on a small patient sample and should therefore be considered exploratory. However, this study also has several key strengths. It is a large national prospective cohort study, which enables correlations to be investigated between viral genotype and clinical phenotype in critically ill patients.

## Conclusions

This French national prospective observational study, found that the emergence of the influenza A(H3N2) subclade K was associated with an increased risk of death in univariable but not multivariable analysis, adjusting for host-related factors.

## ABBREVIATIONS

AHRF: acute hypoxemic respiratory failure
HA: hemagglutinin
ICU: Intensive care unit
IMV: Invasive mechanical ventilation

## DECLARATION

### ETHICAL APPROVAL

The study was approved by the Comité de Protection des Personnes Sud-Méditerranée I (N° EudraCT/ID-RCB: 2021-A02914-37). Informed consent was obtained from all patients or their relatives. The study was conducted in accordance with the 1964 Helsinki Declaration and subsequent amendments.

## DATA AVAIBILITY

the clinical datasets generated during and/or analyzed during the current study are available from the corresponding author on reasonable request (N.D.P.).

## ACKNOWLEDGMENTS

The authors would like to thank all study investigators, Dr Pierre-André Natella, Ms Nolwenn Bombenger for taking care of regulatory aspects, Ms Clélia Chambraud for taking care of data management, Mr Mohamed Ader for clinical data abstraction, the nurses and physicians who took care of the patients, the laboratory staff who took care of virological samples and the patients and their family for agreeing to participate in the study.

## AUTHOR’S CONTRIBUTIONS

Nicolas de Prost, Etienne Audureau, Jean-Michel Pawlostky and Slim Fourati designed the study and obtained funding; Etienne Audureau performed statistical analyses; Pierre Bay, Nicolas de Prost, Sebastien Préau, François M Beloncle, Anaïs Dartevel, Arnaud W. Thille, Cédric Darreau, Cécile Aubron, Damien Roux, Fabienne Tamion, Antoine Guillon, Bertrand Souweine, Kada Klouche and Charles Damoisel, included the patient and were responsible for clinical data collection; Aurélie Guigon, Caroline Lefeuvre, Sylvie Larrat, Luc Deroche, Jean Thomin, Adissa Tran, Quentin Le Hingrat, Alice Moisan, Lynda Handala, Cecile Henquell, Edouard Tuaillon, Anne-Marie Roque-Afonso, Alexandre Soulier, and Slim Fourati were responsible of the management of virological samples; Jean-Michel Pawlostky, Alexandre Soulier, Marie Le Goff, Frédéric Lemoine, Marie-Anne Rameix-Welti and Slim Fourati were responsible of virological analyses; Pierre Bay, Nicolas de Prost, Marie Le Goff, Etienne Audureau, and Slim Fourati wrote the first draft of the article; all authors revised and approved the article. the corresponding author attests that all listed authors meet authorship criteria and that no others meeting the criteria have been omitted. Nicolas de Prost is the guarantor.

## FINANCIAL/NONFINANCIAL DISCLOSURES

Slim Fourati has served as a speaker for GlaxoSmithKline, AstraZeneca, MSD, Pfeizer, Cepheid and Moderna, SANOFI. Jean-Michel Pawlotsky has served as an advisor or speaker for Abbvie, Arbutus, Assembly Biosciences, Gilead and Merck. Etienne Audureau has received fees for lectures from Alexion, Sanofi, Gilead and Pfizer. His hospital has received research grant from Pfizer, MSD and Alexion. All others authors have nothing to disclose. Rémi COUDROY received fees and reimbursement for travel expenses from Fisher and Paykel Healthcare. Sylvie LARRAT received reimbursement for travel expenses from GILEAD and Biomerieux.

## ROLE OF THE SPONSORS

The SEVARVIR study has been funded by the EMERGEN consortium—ANRS Maladies Infectieuses Emergentes (ANRS0153). This study has been labeled as a National Research Priority by the National Orientation Committee for Therapeutic Trials and other researches on Covid-19 (CAPNET). The investigators would like to acknowledge ANRS | Emerging infectious diseases for their scientific support, the French Ministry of Health and Prevention and the French Ministry of Higher Education, Research and Innovation for their funding and support.

## OTHER CONTRIBUTIONS

None.

## MEDICAL WRITING, EDITORIAL AND OTHER ASSISTANCE

none

## CONSORTIA - THE SEVARVIR INVESTIGATORS

Henri Mondor, Créteil, Medical ICU: Nicolas DE PROST, Pierre BAY, Keyvan RAZAZI, Armand MEKONTSO DESSAP; Henri Mondor, Créteil, Surgical ICU: Lucile PICARD, Nicolas MONGARDON; Henri Mondor Créteil, Virology: Slim FOURATI; Alexandre SOULIER; Mélissa N’DEBI; Sarah SENG; Mohamed ADER; Pierre CAPPY; Christophe RODRIGUEZ; Jean-Michel PAWLOTSKY; Henri Mondor, Créteil, URC: Etienne AUDUREAU, Pierre-André NATELLA; Cochin, Paris, Medical ICU: Frédéric PENE; Cochin, Paris, Virology: Anne-Sophie L’HONNEUR_; Saint-Louis, Paris, Medical ICU: Adrien JOSEPH, Elie AZOULAY, Megan FRAISSE; Saint-Louis, Paris, Virology: Maud SALMONA; Marie-Laure CHAIX_; Pitié-Salpêtrière, Paris, Medical and cardiac ICU: Charles-Edouard LUYT, David LEVY; Pitié-Salpêtrière, Paris, Medical ICU: Julien MAYAUX_; Pitié-Salpêtrière, Paris, Virology_: Stéphane MAROT, Saint-Antoine, Paris, Medical ICU: Juliette BERNIER; Vincent BONNY, Tomas URBINA, Hafid AIT-OUFELLA, Eric MAURY; Saint-Antoine, Paris, Virology_: Laurence MORAND-JOUBERT; Djeneba BOCAR FOFANA; Bichat, Paris, Medical ICU: Jean-François TIMSIT; Bichat, Paris, Virology: Diane DESCAMPS, Quentin LE HINGRAT; Tenon, Paris, Medical and Surgical ICU: Guillaume VOIRIOT, Nina DE MONTMOLLIN, Mathieu TURPIN; Tenon, Paris, Virology: Laurence MORAND-JOUBERT, Avicenne, Bobigny, Medical and Surgical ICU_: Stéphane GAUDRY; Avicenne, Bobigny, Virology: Ségolène BRICHLER; Louis Mourier, Colombes, Medical and Surgical ICU: Fabrice Uhel, Damien Roux; Microbiology, Luce LANDRAUD, Anne-Claire MAHERAULT, Bicêtre, Le Kremlin-Bicêtre, Medical ICU: Tài Olivier PHAM; Bicêtre, Le Kremlin-Bicêtre, Virology: Amal CHAGHOURI, Anne-Marie ROQUE-ALFONSO; Raymond Poincaré, Garches, Medical ICU_: Nicholas Heming, Djillali Annane, Marie-Alice BOVY; Ambroise Paré, Boulogne, Medical and Surgical ICU: Sylvie Meireles, Antoine Vieillard-Baron; Ambroise Paré, Boulogne, Virology: Elyanne GAULT; Hôpital Marc Jacquet, Melun, Medical ICU: Sébastien Jochmans; Hôpital Marc Jacquet, Melun, Microbiology: Aurélia PITSCH; CH Sud Francilien, Corbeil-Essonnes, ICU: Guillaume CHEVREL, Céline CLERGUE; CH Sud Francilien, Corbeil Essonnes, Microbiology: Kubab SABAH; CH Victor Dupouy, Argenteuil, ICU: Damien CONTOU; CH Victor Dupouy, Argenteuil, Microbiology: Amandine HENRY; Saint-Camille, Bry-sur-Marne, Polyvalent ICU: Malo EMERY; Saint-Camille, Bry-sur-Marne, Microbiology: Claudio GARCIA-SANCHEZ; CHU de Strasbourg, Medical ICU: Ferhat MEZIANI; Louis-Marie JANDEAUX; CHU de Strasbourg, Virology: Samira FAFI-KREMER; Elodie LAUGEL; CHU de Lille, Medical ICU: Sébastien PREAU, Raphaël Favory, Claire BOUREL, Côme BUREAU, Estelle DANCHE, Alexandre GAUDET, Geoffrey LEDOUX; CHU de Lille, Virology, Aurélie GUIGON; CHRU de Nancy, Hôpitaux de Brabois, Medical ICU: Antoine KIMMOUN_; CHRU de Nancy, Hôpitaux de Brabois, Virology: Evelyne SCHVOERER, Cédric HARTARD; Antoine Béclère, Clamart, General ICU: Charles DAMOISEL; Antoine Béclère, Clamart, Virology_: Anne-Marie Roque-Alfonso, Hôpital Européen Georges Pompidou, Paris, Medical ICU: Nicolas BRECHOT; Hôpital Européen Georges Pompidou, Paris, Virology: Hélène PÉRÉ; CHU Tours, Medical ICU: Antoine GUILLON; CHU Tours, Virology_: Lynda Handala_; CHU d’Angers, Medical ICU: François BELONCLE, Achille KOUACHET, Mathilde TAILLANTOU-CANDAU; CHU d’Angers, Virology_: Caroline LEFEUVRE_; Alexandra DUCANCELLE; CHU de Poitiers, Medical ICU_: Rémi COUDROY, Arnaud W. THILLE, François ARRIVE, Sylvain LE PAPE, Laura MARCHASSON; CHU de Poitiers, Virology: Luc DEROCHE, Nicolas LEVEQUE; CHU de Rennes, Medical ICU: Jean-Marc Tadié, –Valentin COIRIER; CHU de Rennes, Virology_: Vincent THIBAULT; Claire GROLHIER; CH de Lorient, Medical ICU: Béatrice LA COMBE; CH de Lorient, Microbiology_: Séverine HAOUISEE; CHU de Bordeaux, Medical ICU: Alexandre BOYER_; CHU de Bordeaux, Virology: Sonia BURREL; CHU de Rouen, Medical ICU: Fabienne TAMION; Gaetan BEDUNEAU, Christophe GIRAULT, Maximillien GRALL, Dorothée CARPENTIER; CHU de Rouen, Virology_: Alice, MOISAN; Elodie Alessandri-Gradt; Véronique Lemée; Adeline Baron; Marie Gueudin; CHU de Nantes, Medical ICU_: Emmanuel CANET; CHU de Nantes, Virology_: Audrey, RODALLEC, Berthe Marie IMBERT; CHU de Nice, Medical ICU: Clément SACCHERI; CHU de Nice, Virology: Géraldine GONFRIER; Marseille Hôpital Nord, Medical ICU: Sami HRAEICH_; Marseille Hôpital Nord, IHU Méditerranée: Pierre-Edouard FOURNIER, Philippe COLSON; CH Le Mans, General ICU: Cédric Darreau; CH Le Mans, Microbiology: Jean THOMIN; CHU de Grenoble, Medical ICU: Anaïs DARTEVEL; CHU de Grenoble, Virology: Sylvie LARRAT; Raphaële GERMI; Aurélie TRUFFOT_ CHU de Saint-Etienne, Medical ICU: Guillaume THIERY; CHU de Saint-Etienne, Virology_: Sylvie PILLET; CHU de Montpellier, Medical ICU: Kada KLOUCHE; CHU de Montpellier, Virology_: Edouard TUAILLON; CHU de Brest, Medical ICU: Cécile AUBRON; CHU de Brest, Virology_: Adissa TRAN, Sophie VALLET; CHU de Dijon, Medical ICU: Pierre-Emmanuel CHARLES; CHU de Dijon, Virology: Alexis DE ROUGEMONT; CHU de Clermont-Ferrand, Medical ICU: Bertrand SOUWEINE; CHU de Clermont-Ferrand, Virology_: Cecile HENQUELL; Audrey MIRAND; CHU de Reims, Medical ICU: Bruno MOURVILLIER_; CHU de Reims, Virology: Laurent ANDREOLETTI, Quentin LECOMTE-THENOT; CHU de Caen, Medical ICU_: Damien DU CHEYRON; CHU de Caen, Virology_: Nefert CANDACE DOSSOU; Astrid VABRET; CHU de Besançon, Medical ICU: Gaël PITON, Jean-Christophe NAVELLOU; CHU de Besançon, Virology: Quentin LEPILLER; CHU de Limoges, Medical ICU: Thomas DAIX; CHU de Limoges, Virology: Sébastien HANTZ, Sylvie ROGER, CHU de Toulouse, Medical ICU_: Benjamine SARTON, CHU de Toulouse, Virology_: Camille VELLAS, CHU de Nîmes, Medical ICU_: Claire ROGER, CHU de Nîmes, Virology_: Robin STEPHAN, CH Metz-Thionville, Medical ICU_: Guillaume LOUIS, CH Metz-Thionville, Virology_: Christelle JOST, Pascale PEREZ, CHU Nancy-Hôpital Central, Medical ICU_: Sébastien GIBOT, CHU Nancy-Hôpital Central, Virology_: Evelyne SCHVOERER, CHU Orléans, Medical ICU_: François BARBIER, CHU Orléans, Virology_: Dr Jerome Guinard / Pr Véronique Avettand-Fenoel, CH Dieppe, Medical ICU_: Jean-Philippe RIGAUD, CH Dieppe, Virology_: Elodie BLONDEL, CHU Réunion – Saint-Denis, Medical ICU_: Amélie RENOU, CHU Réunion – Saint-Denis, Virology_: Laurent SOUPLY, CHU Martinique, Medical ICU_: Hossein MEHDAOUI, CHU Martinique, Virology_: Laurence FAGOUR, CH de Tourcoing, Medical ICU_: Hugues GEORGES, CH de Tourcoing, Virology_: Pierre PATOZ, CH de Haguenau, Medical ICU_: Asaël BERGER, CH de Haguenau, Virology_: Julien EXINGER, Hospices Civils de Lyon, Medical ICU_: Jean-Christophe RICHARD, Hospices Civils de Lyon, Virology_: Alexandre GAYMARD, CHU de Strasbourg, Infectious Diseases_: François DANION, CHU de Rouen, Infectious Diseases_: Manuel ETIENNE, CHU de Grenoble, Infectious Diseases_: Anne-Laure MOUNAYAR, CHU de Nice, Infectious Diseases_: Matthieu BUSCOT, CHU de Rennes, Infectious Diseases_: David LUQUE-PAZ, CHU de Nîmes, Infectious Disease_: Paul LOUBET, Romaric LARCHER, Myriam CHIARUZZI, Aurélie MARTIN, Marie POUPARD, Fanny VILLA, CHU de Toulouse, Infectious Diseases_: Guillaume MARTIN-BLONDEL, Hospices Civils de Lyon, Infectious Diseases_: Florence ADER, CHU de Lille, Infectious Diseases_: Emmanuel FAURE, Henri Mondor, Créteil, Infectious Diseases_: Giovanna MELICA, Bichat, Paris, Infectious Diseases_: Nathan PEIFFER-SMADJA, Raymond Poincaré, Garches, Infectious Disease_: Aurélien DINH, Saint-Antoine, Paris, Infectious Diseases, Karine LACOMBE

## Notes

### Clinical Protocols

https://clinicaltrials.gov/study/NCT05162508

### Author Declarations

The study was approved by the Comite de Protection des Personnes Sud-Mediterranee I (Number EudraCT/ID-RCB: 2021-A02914-37). Informed consent was obtained from all patients or their relatives. The study was conducted in accordance with the 1964 Helsinki Declaration and subsequent amendments.

