## Supplementary material for "ICU patients with influenza during the 2025–26 season: a French prospective nationwide cohort study"

**AUTHORS:** Nicolas de Prost^1,2,3^, Pierre Bay^1,2,3^, Marie Le Goff^1,2,3^, Sébastien Préau^4,5^, Aurélie Guigon^6^, François M Beloncle^7^, Caroline Lefeuvre^8^, Anaïs Dartevel^9^, Sylvie Larrat^10^, Rémi Coudroy^11,12^, Luc Deroche^13,14^, Cédric Darreau^15^, Jean Thomin^16^, Cécile Aubron^17^, Adissa Tran^18^, Fabrice Uhel^19,20^, Quentin Le Hingrat^21^, Fabienne Tamion^22^, Alice Moisan^23^, Antoine Guillon^24^, Lynda Handala^25^, Bertrand Souweine^26^, Cecile Henquell^27^, Kada Klouche^28,29^, Edouard Tuaillon^30,31^, Charles Damoisel^32^, Anne-Marie Roque-Afonso^33^, Elyanne Gault^34^, Pierre Cappy^3,35^, Alexandre Soulier^3,35^, Jean-Michel Pawlotsky^3,35^, Frédéric Lemoine^†36^, Marie-Anne Rameix-Welti^†37^, Etienne Audureau*^38^, Slim Fourati*^3,35^, on behalf of the SEVARVIR investigators

† contributed equally

* contributed equally as last authors

#### Methods

### Influenza sequencing

Full-length influenza A and B virus genomes (8 segments) were obtained using a whole-genome amplification strategy based on the French National Influenza Reference Center protocol. Viral RNA was extracted from respiratory specimens using the QIAsymphony DSP Virus/Pathogen Kit on a QIAsymphony automated platform (Qiagen, Hilden, Germany), according to the manufacturer’s instructions. Reverse transcription and multiplex PCR amplification of the eight genomic segments were performed using a one-step RT-PCR approach with segment-specific universal primers targeting the conserved 5′ and 3′ terminal regions of influenza A or B viruses. Thermal cycling included reverse transcription, pre-amplification, and 35 amplification cycles. Sequencing libraries were prepared using a Nextera-based workflow (Illumina, San Diego, CA, USA) and sequenced on a NovaSeq platform (Illumina) according to the manufacturer’s instructions. Demultiplexed reads were assembled to generate consensus sequences for each of the eight genomic segments.

### Phylogenetic analyses

We randomly selected 300 high-quality French contextual samples of each subtype from the NRC (spanning the 2024–2025 and 2025–2026 seasons) using goalign subset (version devd0f3e37) ^1^. SEVARVIR sequences containing >20% ambiguous nucleotides (N) were filtered out using goalign clean ^1^, and those derived from samples with high CT values (≥30) were excluded. All sequences were annotated with Nextclade v3.18.0 ^2^ and aligned using Augur v32.1.0 ^3^. Maximum-likelihood phylogenetic trees were inferred with IQTree v3.0.1 ^4^ under the GTR+I+G4 substitution model (specifying --ninit 20 --epsilon 0.0). Ancestral nucleotide and amino acid sequences were reconstructed using Augur (refine, ancestral, and translate commands) ^3^. Final trees were converted to Newick format with gotree ^1^ and visualized using iTOL ^5^.

### Statistical analyses

To illustrate differences in phenotypes of patients according to infecting influenza virus subtype status, we performed an exploratory unsupervised clustering analysis using the Kohonen’s self-organized map (SOM) methodology ^6^, allowing us to build 2-dimensional maps from multidimensional datasets. In a nutshell, each map is divided into districts in which patients are located by the SOM algorithm on the basis of their characteristics: patients with similar features are closely located on the maps, while patients with distinct profiles are farther from each other, hence allowing to identify key differences or similarities among them. The SOMs were obtained with the Numero package framework for the R statistical platform ^7^ after variables with missing information were imputed using the k-nearest neighbors (k-NN) approach and principal component analysis adapted for mixtures of qualitative and quantitative variables was applied (PCAMix) ^8,9^. All measurements were taken from distinct samples. Analyses were performed using Stata V16.1 statistical software (StataCorp, College Station, TX, USA), and R 4.2.0 (R Foundation for Statistical Computing, Vienna, Austria).

#### eTable 1. List of centres participating in the SEVARVIR study

| n° | Centre | Department |
| --- | --- | --- |
| 001 | Henri Mondor, Créteil | Medical ICU |
| 002 | Henri Mondor, Créteil | Surgical ICU |
| 003 | Cochin, Paris | Medical ICU |
| 004 | Saint-Louis, Paris | Medical ICU |
| 005 | Pitié-Salpêtrière, Paris | Medical ICU |
| 006 | Saint-Antoine, Paris | Medical ICU |
| 007 | Pitié-Salpêtrière, Paris | Pulmonary and Medical ICU |
| 008 | Bichat, Paris | Medical ICU |
| 009 | Tenon, Paris | ICU |
| 010 | Avicenne, Bobigny | ICU |
| 011 | Louis Mourier, Colombes | ICU |
| 012 | Bicêtre, Le Kremlin-Bicêtre | Medical ICU |
| 013 | Raymond Poincaré, Garches | Medical ICU |
| 014 | Ambroise Paré, Boulogne-Billancourt | ICU |
| 015 | Marc Jacquet, Melun | ICU |
| 016 | CH Sud Francilien, Jossigny | ICU |
| 017 | CH Victor Dupouy, Argenteuil | ICU |
| 018 | Saint Camille, Bry sur Marne | ICU |
| 019 | CHU de Strasbourg | ICU |
| 020 | CHU de Lille | ICU |
| 021 | CHRU de Nancy, Hôpitaux de Brabois - Nancy | Medical ICU |
| 022 | Antoine Béclère, Le Plessy Robinson | ICU |
| 023 | HEGP, Paris | Medical ICU |
| 024 | CHU de Tours | Medical ICU |
| 025 | CHU de Poitiers | Medical ICU |
| 026 | CHU de Rennes | Medical ICU |
| 027 | CH de Lorient | Medical ICU |
| 028 | CHU de Rouen | Medical ICU |
| 029 | CHU de Nantes | Medical ICU |
| 030 | CHU de Nice | Medical ICU |
| 031 | CH du Mans | ICU |
| 032 | CHU de Grenoble | Medical ICU |
| 033 | CHU de Saint-Etienne | Medical ICU |
| 034 | CHU de Montpellier | Medical ICU |
| 035 | CHU de Brest | Medical ICU |
| 036 | CHU de Clermont-Ferrand | Medical ICU |
| 037 | CHU de Reims | Medical ICU |
| 038 | CHU de Caen | Medical ICU |
| 039 | CHU de Limoges | Medical ICU |
| 040 | CHR Metz-Thionville - Hôpital Mercy | ICU |
| 041 | CH de Dieppe | ICU |
| 042 | CH de Tourcoing | ICU |

| eTable 2. Characteristics of the 325 critically ill patients infected with influenza according to the infecting subtype during 2025-2026 season (n=325) | | | | | | |
| --- | --- | --- | --- | --- | --- | --- |
| **Variable** | **N** | **H1N1  N=70** | **H3N2  N=74** | **Subtype undetermined**  **N=34** | **Unavailable for sequencing**  **N=147** | **p** |
| **Demographics and comorbidities** | | | | | | |
| Sex, females | 325 | 27 (38.6) | 29 (39.2) | 14 (41.2) | 60 (40.8) | 0.99 |
| Age, years | 325 | 66.6 [56.5;74.7] | 72.0 [61.6;77.6] | 67.9 [56.5;73.7] | 67.3 [59.0;73.8] | 0.12 |
| Diabetes | 230 | 18 (34.0) | 24 (38.1) | 7 (28.0) | 26 (29.2) | 0.67 |
| Obesity | 318 | 26 (38.2) | 19 (26.4) | 6 (17.6) | 40 (27.8) | 0.22 |
| Chronic heart failure | 229 | 8 (15.4) | 12 (19.0) | 4 (16.0) | 16 (18.0) | 0.97 |
| Hypertension | 230 | 24 (45.3) | 44 (69.8) | 11 (44.0) | 46 (51.7) | **0.025** |
| Chronic respiratory failure | 230 | 17 (32.1) | 23 (36.5) | 12 (48.0) | 38 (42.7) | 0.46 |
| Chronic renal failure | 230 | 7 (13.2) | 13 (20.6) | 2 (8.0) | 9 (10.1) | 0.27 |
| Cirrhosis | 230 | 0 (0.0) | 1 (1.6) | 0 (0.0) | 0 (0.0) | 0.61 |
| Immunosuppression | 230 | 15 (28.3) | 9 (14.3) | 5 (20.0) | 17 (19.1) | 0.31 |
| Immunosuppression category | 220 |  | | | | 0.75 |
| None |  | 38 (77.6) | 54 (88.5) | 20 (87.0) | 72 (82.8) |  |
| Solid organ transplant |  | 1 (2.0) | 3 (4.9) | 0 (0.0) | 3 (3.4) |  |
| Onco-hematological malignancies |  | 5 (10.2) | 2 (3.3) | 1 (4.3) | 6 (6.9) |  |
| Others^1^ |  | 5 (10.2) | 2 (3.3) | 2 (8.7) | 6 (6.9) |  |
| Number of comorbidities | 230 | 2.0 [1.0;2.0] | 2.0 [1.0;3.0] | 2.0 [1.0;3.0] | 2.0 [1.0;3.0] | 0.33 |
| Clinical frailty scale | 265 | 3.0 [1.0;4.0] | 3.0 [2.0;4.0] | 2.0 [1.0;4.0] | 3.0 [2.0;4.0] | 0.61 |
| **Influenza infection and Vaccination** | | | | | | |
| Previous Influenza infection | 209 | 3 (6.4) | 13 (23.2) | 2 (8.7) | 14 (16.9) | 0.095 |
| Influenza vaccination | 218 | 18 (34.0) | 22 (36.1) | 6 (23.1) | 16 (20.5) | 0.15 |
| Influenza RNA detection in nasopharyngeal swabs, Ct | 178 | 30.0 [25.0;34.0] | 30.0 [25.0;35.0] | 40.0 [40.0;40.0] | NA | **<0.001** |
| **Patients severity upon ICU admission and biological features** | | | | | | |
| SAPS II score | 256 | 39.5 [29.0;49.0] | 35.0 [25.0;49.0] | 36.5 [27.5;50.5] | 39.0 [31.0;51.0] | 0.71 |
| SOFA score | 275 | 5.0 [3.0;8.5] | 4.0 [3.0;8.0] | 4.0 [3.0;6.0] | 5.0 [2.0;9.0] | 0.63 |
| PaO2/FiO2 ratio, mmHg | 267 | 139 [98;223] | 197 [115;280] | 140 [97;228] | 163 [108;241] | 0.39 |
| Arterial lactate level, mM | 266 | 1.50 [1.10;2.45] | 1.40 [1.00;2.40] | 1.70 [1.00;2.60] | 1.60 [1.20;2.40] | 0.45 |
| Blood leukocytes, G/L | 286 | 8.7 [4.50;13.3] | 8.8 [5.2;10.9] | 8.9 [7.0;11.2] | 9.4 [6.1;13.6] | 0.52 |
| Blood lymphocytes, G/L | 243 | 0.60 [0.40;1.00] | 0.60 [0.40;1.05] | 0.60 [0.40;1.60] | 0.80 [0.40;1.20] | 0.48 |
| Blood platelets, G/L | 286 | 158 [118;242] | 190 [145;231] | 215 [135;263] | 205 [145;268] | 0.051 |
| Serum urea level, mM | 285 | 7.0 [5.0;13.8] | 8.3 [5.2;12.6] | 5.7 [3.50;8.2] | 7.4 [5.2;12.3] | 0.076 |
| Serum creatinine level, µM | 287 | 94.0 [58.5;146] | 89.0 [65.0;141] | 74.0 [58.0;93.0] | 81.0 [61.0;133] | 0.31 |
| *Oxygen/ventilatory support* | 285 |  | | | | 0.67 |
| Oxygen |  | 15 (22.1) | 19 (25.7) | 5 (15.2) | 25 (22.7) |  |
| High flow oxygen |  | 15 (22.1) | 19 (25.7) | 14 (42.4) | 26 (23.6) |  |
| NIV/C-PAP |  | 14 (20.6) | 14 (18.9) | 4 (12.1) | 23 (20.9) |  |
| Invasive MV |  | 24 (35.3) | 22 (29.7) | 10 (30.3) | 36 (32.7) |  |
| Vasopressor support | 285 | 24 (35.3) | 22 (29.7) | 10 (30.3) | 36 (32.7) | 0.90 |
| Bacterial coinfection | 282 | 15 (22.1) | 17 (23.3) | 10 (31.3) | 40 (36.7) | 0.12 |
| **Management during ICU stay** | | | | | | |
| Invasive MV | 287 | 28 (41.2) | 22 (29.7) | 11 (33.3) | 45 (40.2) | 0.42 |
| Prone positioning | 261 | 16 (26.2) | 6 (8.2) | 3 (9.7) | 16 (16.7) | **0.028** |
| MV duration, days* | 68 | 12.0 [9.0;20.0] | 7.0 [3.0;17.0] | 10.0 [7.0;17.0] | 8.0 [5.0;14.0] | 0.34 |
| Live-ventilator-free days at day-28* | 171 | 28.0 [16.0;28.0] | 28.0 [11.0;28.0] | 28.0 [18.0;28.0] | 26.0 [12.0;28.0] | 0.56 |
| Vasopressor support | 284 | 24 (35.8) | 21 (28.4) | 11 (33.3) | 38 (34.5) | 0.78 |
| Duration of vasopressors, days* | 68 | 4.0 [3.0;10.0] | 3.0 [1.0;10.0] | 5.0 [2.0;13.0] | 2.50 [1.0;8.0] | 0.35 |
| Renal Replacement Therapy | 280 | 11 (16.4) | 5 (6.8) | 3 (9.1) | 12 (11.3) | 0.32 |
| Ventilator-acquired pneumonia* ^2^ | 205 | 8 (13.1) | 3 (4.4) | 2 (7.1) | 7 (14.6) | 0.20 |
| Oseltamivir | 245 | 53 (93.0) | 62 (95.4) | 30 (93.8) | 85 (93.4) | 0.96 |
| Corticosteroids | 243 | 15 (26.3) | 21 (32.8) | 13 (40.6) | 32 (35.6) | 0.52 |
| **Outcomes** | | | | | | |
| Day-28 mortality* | 170 | 4 (7.8) | 11 (20.0) | 4 (16.7) | 9 (22.5) | 0.19 |
| Duration of ICU stay, days* | 201 | 7.0 [4.0;16.0] | 5.0 [3.0;9.0] | 5.0 [4.0;10.0] | 6.0 [3.0;11.5] | 0.30 |
| Results are N (%), means (±standard deviation) or medians (interquartile range, i.e., quartile 1; quartile 3).  ICU intensive care unit, Ct cycle threshold, WHO World Health Organization, SOFA Sequential Organ Failure Assessment, SAPS II Simplified Acute Physiology Score II, NIV non-invasive ventilation, C-PAP continuous-positive airway pressure, MV mechanical ventilation.  ^1^Includes HIV infection, long-term corticosteroid treatment, and other immunosuppressive treatments.  ^2^VAP episodes were recorded per definition in patients under IMV since more than 48 h.  *These data are reported only for patients with available follow-up of at least 30 days (H1N1: *N*=62; H3N2 season: *N*=68; subtype undetermined *N*=29; unavailable for sequencing *N*=50).  Two-tailed p-values p-values come from unadjusted comparisons using Chi-square or Fisher’s exact tests for categorical variables, and Kruskal-Wallis tests for continuous variables, as appropriate. No adjustment for multiple comparisons was performed. Bolded p-values are significant at the p < 05 level. | | | | | | |

| eTable 3. Independent predictors of mortality by multivariable Cox proportional hazards regression in the critically ill patients infected with influenza during 2025-2026 season (n=158) | | | | | | | |
| --- | --- | --- | --- | --- | --- | --- | --- |
|  | **Univariable Analysis** | | |  | **Multivariable Analysis** | | |
| **Factor** | **HR** | **95%CI** | **p-value** |  | **HR** | **95%CI** | **p-value** |
| *Influenza subtype* |  |  |  |  |  |  |  |
| H1N1 | 1 (ref) |  |  |  | 1 (ref) |  |  |
| H3N2 | 3.24 | (1.05 ; 12.0) | **0.041** |  | 1.13 | (0.32 ; 4.51) | 0.85 |
| Undetermined subtype | 2.91 | (0.69 ; 12.4) | 0.14 |  | 2.42 | (0.33, 13.0) | 0.35 |
| Influenza RNA detection in nasopharyngeal swabs, Ct | 1.00 | (0.94 ; 1.08) | 0.93 |  |  |  |  |
| Sex, females | 0.62 | (0.20 ; 1.67) | 0.35 |  |  |  |  |
| Age, years | 1.06 | (1.01 ; 1.11) | **0.014** |  | 1.05 | (1.00 ; 1.11) | **0.046** |
| Diabetes | 1.05 | (0.36 ; 2.76) | 0.92 |  |  |  |  |
| Obesity | 1.19 | (0.41 ; 3.12) | 0.74 |  |  |  |  |
| Chronic heart failure | 2.17 | (0.69 ; 5.87) | 0.17 |  |  |  |  |
| Hypertension | 5.39 | (1.52 ; 34.2) | **0.006** |  |  |  |  |
| Chronic respiratory failure | 1.46 | (0.52 ; 3.86) | 0.46 |  |  |  |  |
| Chronic renal failure | 0.92 | (0.21 ; 2.83) | 0.89 |  |  |  |  |
| Cirrhosis | 0.00 |  | 0.81 |  |  |  |  |
| Immunosuppression | 1.02 | (0.28 ; 2.90) | 0.98 |  |  |  |  |
| Number of comorbidities | 1.44 | (0.98 ; 2.08) | 0.065 |  |  |  |  |
| Clinical frailty scale | 1.69 | (1.23 ; 2.37) | **0.001** |  | 1.82 | (1.26 ; 2.72) | **0.001** |
| SAPS II score | 1.02 | (0.99 ; 1.04) | 0.14 |  |  |  |  |
| SOFA score | 1.09 | (0.97 ; 1.22) | 0.16 |  |  |  |  |
| PaO2/FiO2 ratio, mmHg | 1.00 | (0.99 ; 1.00) | 0.34 |  |  |  |  |
| Arterial lactate level, mM | 1.00 | (0.79 ; 1.15) | 0.96 |  |  |  |  |
| Blood leukocytes, G/L | 1.02 | (0.95 ; 1.09) | 0.59 |  |  |  |  |
| Blood lymphocytes, G/L | 0.91 | (0.51 ; 1.09) | 0.47 |  |  |  |  |
| Blood platelets, G/L | 1.00 | (1.0 ; 1.00) | 0.91 |  |  |  |  |
| Serum urea level, mM | 1.03 | (0.96 ; 1.09) | 0.36 |  |  |  |  |
| Serum creatinine level, µM | 1.00 | (1.00 ; 1.01) | 0.14 |  |  |  |  |
| Bacterial coinfection | 0.37 | (0.06 ; 1.34) | 0.14 |  |  |  |  |
| Invasive MV upon ICU admission | 2.91 | (1.01 ; 10.5) | **0.049** |  |  |  |  |
| Vasopressor support upon ICU admission | 1.32 | (0.49 ; 3.48) | 0.57 |  |  |  |  |
| ECMO support upon ICU admission | 0.00 |  | 0.30 |  |  |  |  |
| Influenza vaccination | 1.64 | (0.56 ; 4.82) | 0.36 |  |  |  |  |
| Previous Influenza infection | 1.24 | (0.28 ; 3.86) | 0.75 |  |  |  |  |
| Oseltamivir upon ICU admission | 1.80 | (0.41 ; 12.4) | 0.46 |  |  |  |  |
| Corticosteroids upon ICU admission | 0.29 | (0.02 ; 1.62) | 0.18 |  |  |  |  |
| HR (CI 95%): Hazard Ratio (95% confidence interval).  aHR (CI 95%): adjusted Hazard Ratio (95% confidence interval).  *ICU* intensive care unit, *Ct* cycle threshold, ECMO, extracorporeal membrane oxygenation, SOFA Sequential Organ Failure Assessment, *SAPS II* Simplified Acute Physiology Score II, *MV* mechanical ventilation  p-values come from unadjusted Cox proportional hazards regression modeling and multivariable logistic regression models respectively; Bolded p-values are significant at the p < 0.05 level. | | | | | | | |

#### eFigure 1.

Unsupervised analysis of the clinical and biological characteristics of the 158 critically ill patients infected with influenza in 2025-26 with a follow-up period of at least 30 days and a respiratory sample that had been sequenced (*N*=158) by self-organized maps (SOMs). Unsupervised analysis by SOM automatically located patients with similar clinical and paraclinical parameters within 1 of 40 small groupings (“districts”) throughout the map. The closer the patients are on the map, the more similar they are. Each individual map shows the mean values or proportions per district for each characteristic: blue indicates the lowest average values and red indicated the highest, with numbers shown for a selection of representative districts in each SOM. *ICU* intensive care unit, *SOFA* Sequential Organ Failure Assessment, *SAPS II* Simplified Acute Physiology Score II, *MV* mechanical ventilation.

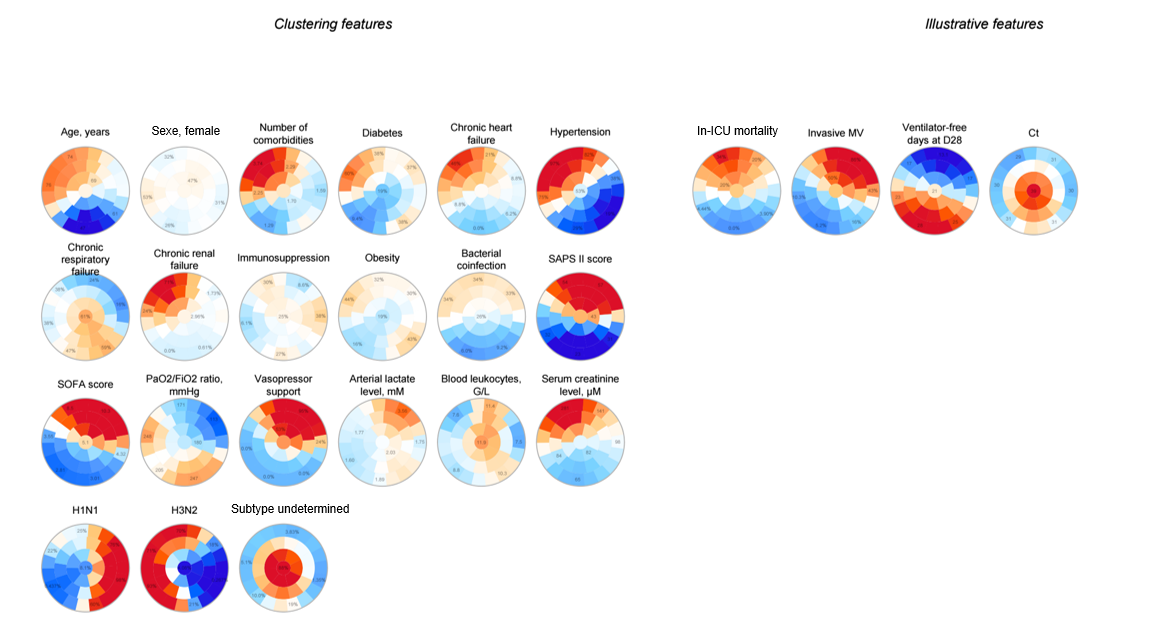
